# A data-driven pipeline to extract potential side effects through co-prescription analysis: application to a cohort study of 2,010 patients taking hydroxychloroquine with an 11-year follow-up

**DOI:** 10.1101/2021.04.14.21255406

**Authors:** P. Sabatier, M. Wack, J. Pouchot, N. Danchin, AS. Jannot

**Affiliations:** Medical Information Department, Georges Pompidou European Hospital, AP-HP, 75015 Paris, France; INSERM U1138, University Paris Descartes, Sorbonne University, 75006 Paris, France; Department of Internal Medicine, Georges Pompidou European Hospital, AP-HP, 75015 Paris, France; Department of Cardiology, Georges Pompidou European Hospital, AP-HP, 75015 Paris, France; University Paris-Descartes, 75006 Paris, France

**Keywords:** chloroquine, hydroxychloroquine, safety, COVID-19, population-wide claim databases

## Abstract

**Context:** Real-life data consist of exhaustive and unbiased data to study drug-safety profiles but are underused because of their complex temporality (i.e., safety depends on the dose, timing, and duration of treatment) and the considerable number of potential side effects to study. We aimed to create a pipeline that manages the complex temporality of real-life data using a data-driven strategy (i.e., without any hypothesis on the potential side effects to search for) to highlight the safety profile of a given drug. We used hydroxychloroquine (HCQ) and its co-prescription in a real-life database to illustrate this pipeline.

**Methods:** We incorporated a weighted cumulative exposure statistical model into a data-driven strategy. This pipeline makes it possible to highlight both long-term and short-term side effects, while avoiding false positives due to the natural course of the underlying disease. We applied the proposed pipeline to a cohort of 2,010 patients with a prescription of HCQ and used their drug prescription as the source of data to highlight the HCQ safety profile.

**Results:** The proposed pipeline introduces a bootstrap strategy into weighted cumulative-exposure statistics estimates to highlight significant drug signals. As applied to HCQ, the proposed pipeline showed nine drugs to be significantly associated with HCQ exposure. Of note, one of them has therapeutic indications for known HCQ side effects. Other associations could be explained by therapeutic indications linked to conditions associated with HCQ indications in France.

**Conclusion:** We propose a data-driven pipeline that makes it possible to provide a broad picture of the side effects of a given drug. It would be informative to pursue the development of this pipeline using other sources of data.

## 1. INTRODUCTION

Adverse drug reactions (ADRs) have been attributed to causing over 770,000 injuries and 100,000 deaths and $76.6 billion in annual costs in the US (1). The rapid detection of ADRs has become a crucial public health issue. Currently, drug safety is monitored by the pharmacovigilance system, which uses spontaneous reporting systems (SRSs) to detect, collect, and analyze ADRs. However, SRSs suffer from underreporting. Indeed, less than 10% of serious ADRs are reported (2,3). Moreover, this system is subject to biases due to selective reporting (most of the reported cases are considered as suspected ADRs).

The increasing availability of electronic healthcare records (EHRs) offers major opportunities to investigate a wide spectrum of ADRs and detect drug safety signals closer to real use and time, as EHR databases record information for large populations over long follow-up periods (4). These databases, such as electronic medical records and administrative claims databases, have been mostly used to confirm or disprove potential signals flagged by SRSs. A number of data-mining techniques have been specifically developed for the automatic detection of drug-safety signals using either SRS or EHR databases (5–11). Over the last decade, several international initiatives have been developed; the Mini-Sentinel and OMOP (Observational Medical Outcomes Partnership) in the United States and the PROTECT (Pharmaco-epidemiological Research on Outcomes of Therapeutics by a European Consortium) and EU-ADR (Exploring and Understanding Adverse Drug Reactions) in Europe.

The challenge of drug-safety signal-detection methods is to handle four types of difficulties. The first difficulty is the data source. The study of long-term adverse drug reactions or effects not suspected by healthcare professionals requires the use of a real-life data source, such as EHR or claims databases, which do not suffer from the known bias of underreporting and reporting selection (12,13). Many drug-safety methods use diagnostic codes from EHR or claims databases to highlight a signal. In most countries, including France, these diagnostic codes are only reported when patients visit hospitals and are therefore strongly biased towards severe side effects. In addition, the diagnostic codes in these databases are primarily reported for financial purposes (reimbursement) and may subsequently be biased towards diagnoses that can be priced. On the contrary, drug reimbursements provide a less biased data source, although they are less precise because a given drug may correspond to several indications and multiple drugs may be prescribed for a given condition. The second difficulty is the consideration of a broad spectrum of potential side effects, and not only candidate side effects (14), to be able to highlight new signals. The third difficulty is to precisely take into account the temporal aspect.

Time is important, because the type of adverse reactions caused by the medication under study may differ according to the duration of the medication prescriptions. Certain adverse effects may occur soon after the start of the medication under study, whereas others may require a prolonged period of administration to become manifest (15). Safety depends on the dose, timing, and duration of the treatment. The last difficulty concerns distinguishing true side effects from the natural course of the disease. Indeed, the natural course of the disease for which the prescribed drug is indicated may be associated with many other diagnoses, which may be indistinguishable from drug side effects using common drug-safety methods.

In this study, we developed a data-driven pipeline based on a cumulative exposure test to highlight the broad picture of side effects of a given drug. We used the French national medication reimbursement database, and given the Covid19-pandemic context, applied this pipeline to HCQ.

## 2. Description of the WCE-AI pipeline

The WCE-AI pipeline aims to overcome the four aforementioned difficulties as follows:

- Data source: We used all drug prescriptions as the unique source of side-effect signals. Drugs are represented at the 5^th^ level of the ATC to group all medications at the active substance level that can be prescribed for the same indication.
- Temporality: We modeled the increase in risk related to cumulative dose using splines in Cox proportional hazards models (16,17). This model is called weighted cumulative exposure (WCE). It allows the representation of complex cumulative effects of dose, timing, and duration of interest drug (18).
- Signal sensitivity: We use bootstrapping to infer the distribution of association statistics between the exposure and each ATC class. This provides insight into the broad spectrum of association signals and highlights those that are significant.
- Specificity of side effects: We introduced a covariate representing disease severity built from expert knowledge to automatically pinpoint drugs given for the condition for which the exposure drug has been prescribed.

We first introduce the WCE-AI pipeline, followed by a presentation of a comparative method to our WCE-AI pipeline. Finally, we present a use case of this pipeline and a comparative method applied to HCQ.

### 2.1 WCE-AI pipeline

The WCE-AI pipeline is divided into three steps: data extraction, data preparation, and safety-profile extraction (Figure 1).

**Figure 1:**
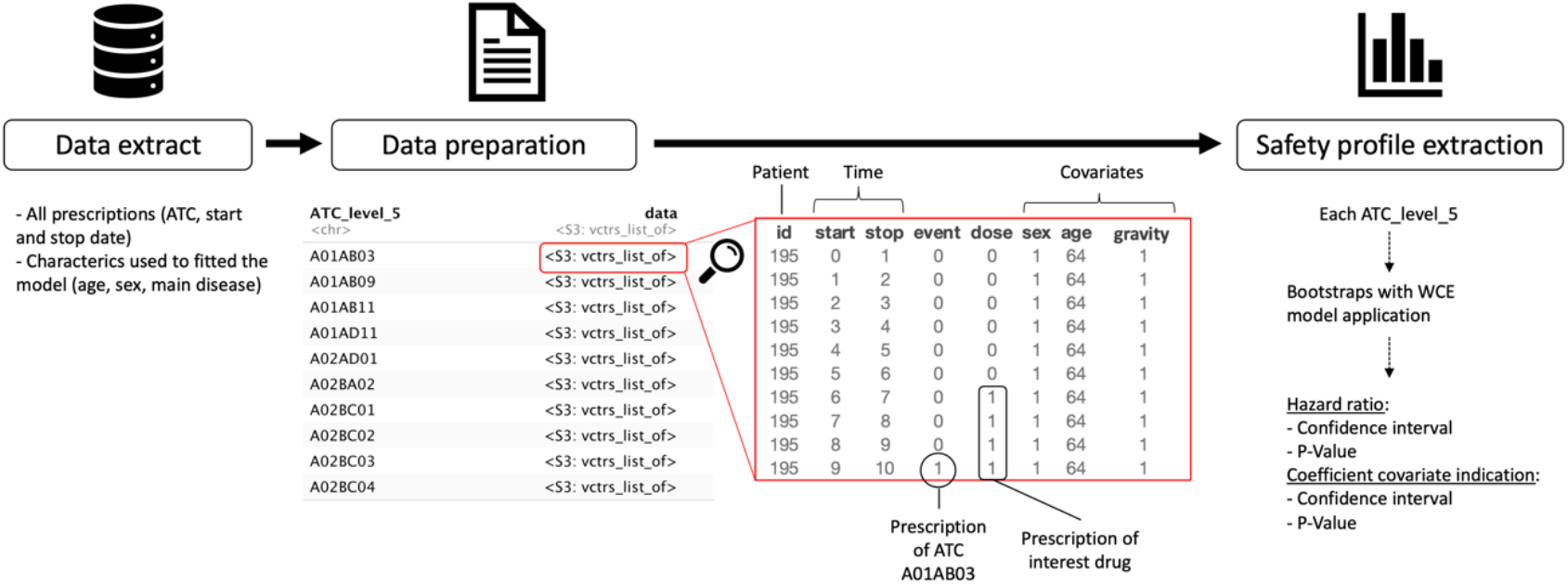
Flow chart of WCE-AI pipeline.

#### i. Data extraction

All drug prescriptions are extracted, along with their treatment initiation date, end of treatment date, ATC class, and patient characteristics, such as age, sex, and disease severity. Disease severity is defined from expert knowledge on the drugs being indicated for the disease for which the exposure is described.

#### ii. Data preparation

We built a data frame corresponding to the WCE package (19) data format for each level 5 ATC class. In each data frame, each row corresponds to a given time period, such as day, week, or month (to be defined). Id identifies the patients. Start and Stop identify the beginning and end of each interval (Stop in row n = Start in row n+1). The intervals are closed on the right. For each patient, the first start (start = 0) is the date of the first drug prescription in the study period. Event is a binary indicator for the event of interest, which has a value of 1 if the event occurred in the interval specified by Start and Stop. For a given subject, event = 1 can only occur in the last interval of his or her follow-up. For each data frame, the event corresponds to the prescription of an level 5ATC class. The last four columns represent three fixed-in-time patient covariates (sex, baseline age, and severity), as well as exposure to the drug of interest. The severity covariate is set to 1 if the patient has been reimbursed at least once for the drug being indicated for the same condition for which it was prescribed (to be defined).

#### iii. Safety-profile extraction

The weighted cumulative exposure (WCE) model is applied to estimate the cumulative effect of duration, dose, and prescription date for the drug of interest (18). The WCE approach estimates the effect of past exposure using a weighted sum of all previous instances of exposure, with weights depending on the time elapsed since exposure and the dose (17,20). This makes it possible to account for the fact that the type of adverse reactions caused by the medication under study may depend on the duration of the medication prescription (certain adverse effects may occur soon after the start of the medication under study, whereas others may require a prolonged period of administration to become manifest). WCE is applied with adjustments for age, sex, and disease severity. For example, to measure the risk of obtaining a prescription of paracetamol/acetaminophen (event) after HCQ exposure (drug of interest), we search for the first prescription of paracetamol/acetaminophen for each patient in the cohort (event). Then, we define potential exposure to HCQ using the WCE method for the six months (time window) before the first paracetamol/acetaminophen prescription (Figure 2).

**Figure 2:**
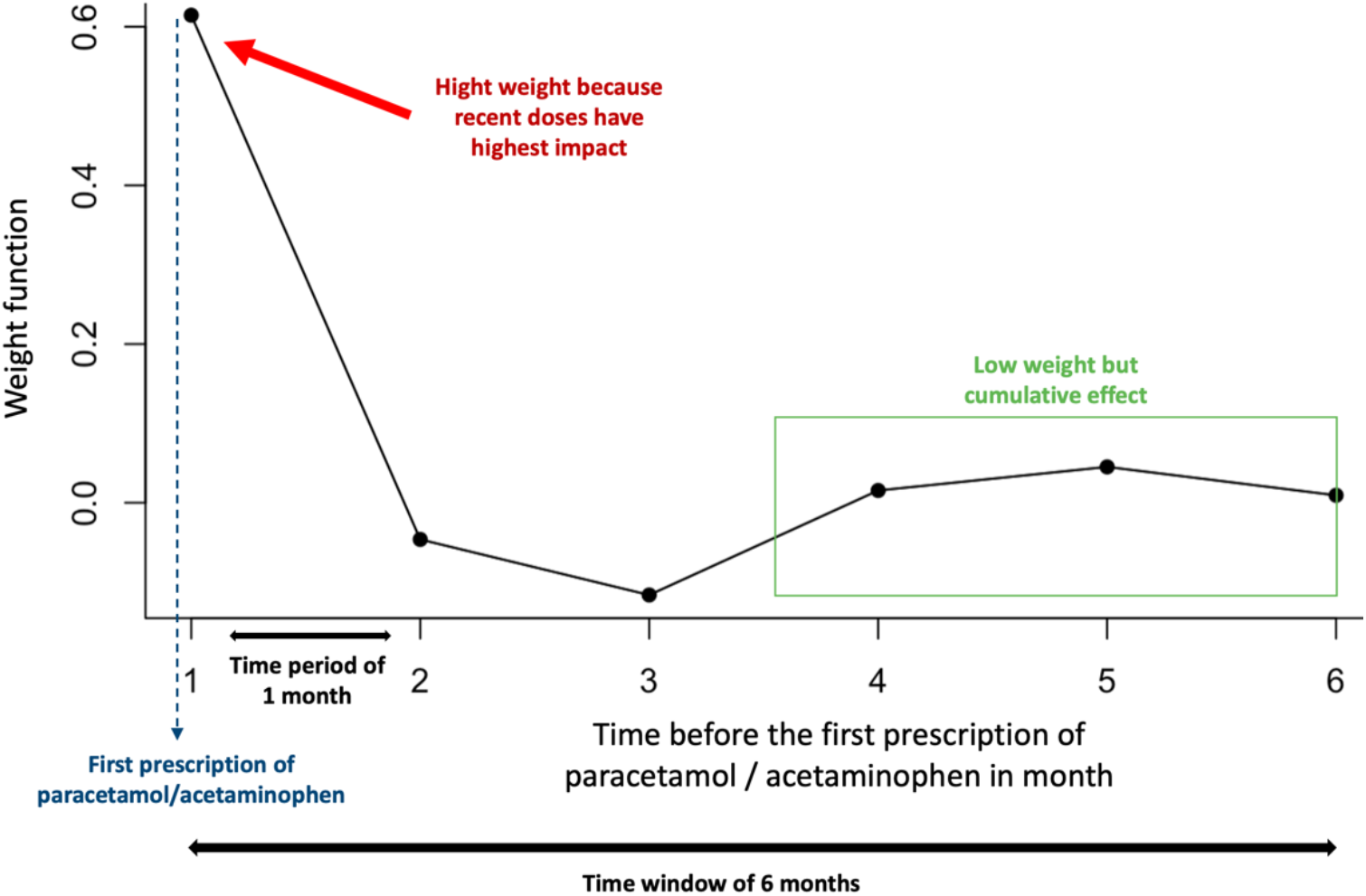
Weight function estimated by the WCE model for a paracetamol prescription after HCQ exposure.

Finally, a Cox proportional hazards model (HR) is fitted to compare a group exposed to the drug of interest (HCQ in this example) during the time window to a group not exposed during the same period.

To show that a drug is significantly associated with exposure, confidence intervals for each hazard ratio are needed. To estimate such confidence intervals, we chose to use the bootstrap method to extract both the confidence intervals and P-values of the HR and the age/sex/severity coefficient (number of repeats to be defined). The bootstrap method is applied for each unique patient identifier. We constructed bootstrap replicates of the data, each of which was randomly sampled with a replacement. Confidence intervals were estimated using the percentile method:

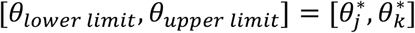

where 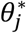 denotes the jth quantile (lower limit) and 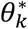 denotes the kth quantile (upper limit)

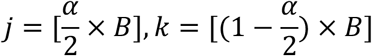

For example, a 95-percentile bootstrap CI with 1,000 bootstrap samples is the interval between the 25th quantile value and the 975th quantile value of the 1,000 bootstrap parameter estimates.

All analyses were performed using R (version 4.0.3) and WCE package (version 1.0.2).

### 2.2 WCE competitor

We compared WCE to self-control case-crossover (SCCO) (21,22), as it relies on the same type of data. SCCO requires to define risk periods and a wash-out period. The case-defining event for each level 5 ATC class was a prescription of a level 5 ATC class other than HCQ. Four periods were defined for each individual with a prescription for the same level 5 ATC class separated by a washout period: one risk period and three control periods. Each period had the same duration (Figure 3). We performed a sensitivity study by varying the length of the periods (risk and control) between 3, 6, and 9 months, with a washout period of one month. The Bonferroni method was used to adjust the P-values to control the type I error rate in multi-testing.

**Figure 3:**
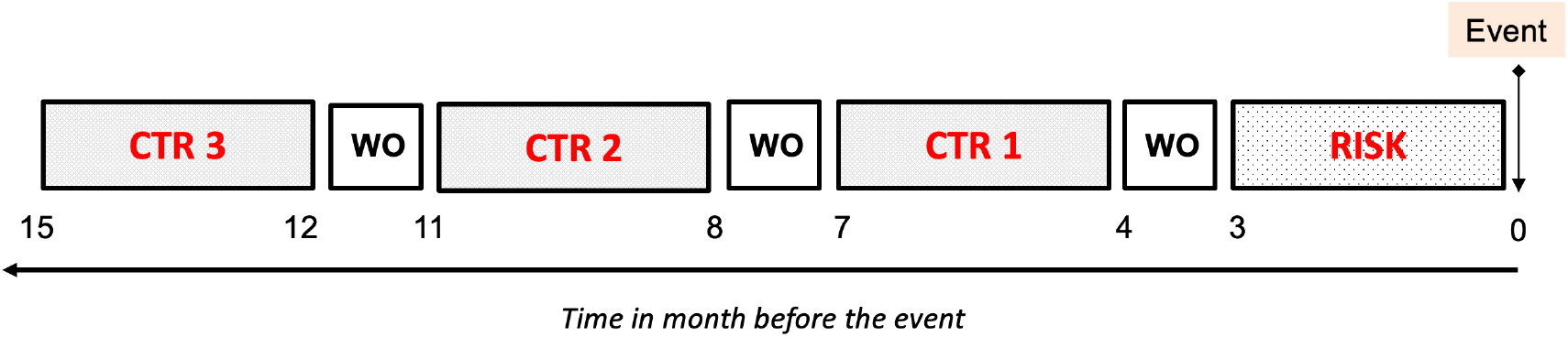
Diagram of self-control case-crossover. Schematic representation of a case–crossover analysis. The case-defining event is the prescription of a level 5 ATC class other than HCQ and the prescription is the HCQ drug. Three control periods are selected (shown as CTR) and one risk period (shown as RISK) for each individual. The duration of the periods (risk and control) is set to three months in this example. The washout period (shown as WO) has a standard duration of one month.

### 2.3. Use case

In the present study, we applied the WCE-AI pipeline to patients newly exposed to HCQ (considered in WCE as the exposure variable). The cohort of patients receiving HCQ was extracted from the EGB, a permanent 1/97 representative sample of the Système National d’Informations Inter-régimes de l’Assurance Maladie (SNIIRAM), which includes the data for 66 million people. The EGB includes data for approximately 780,000 people (23), consisting of de-identified data on demographic characteristics (sex, year of birth, date of death), long-term diseases (ALD), and reimbursed acts (visits, medical procedures, laboratory tests, dispensed drugs, medical devices) (23–25). We extracted the data of all patients with at least one reimbursement for HCQ. We only included patients who did not receive HCQ during the previous year to select only newly exposed patients (Figure 4). For these patients, we extracted the following data: age, sex, date of all drugs deliverances, date of “chronic disease certification status”, which is a marker of disease severity in the French health system.

**Figure 4.**
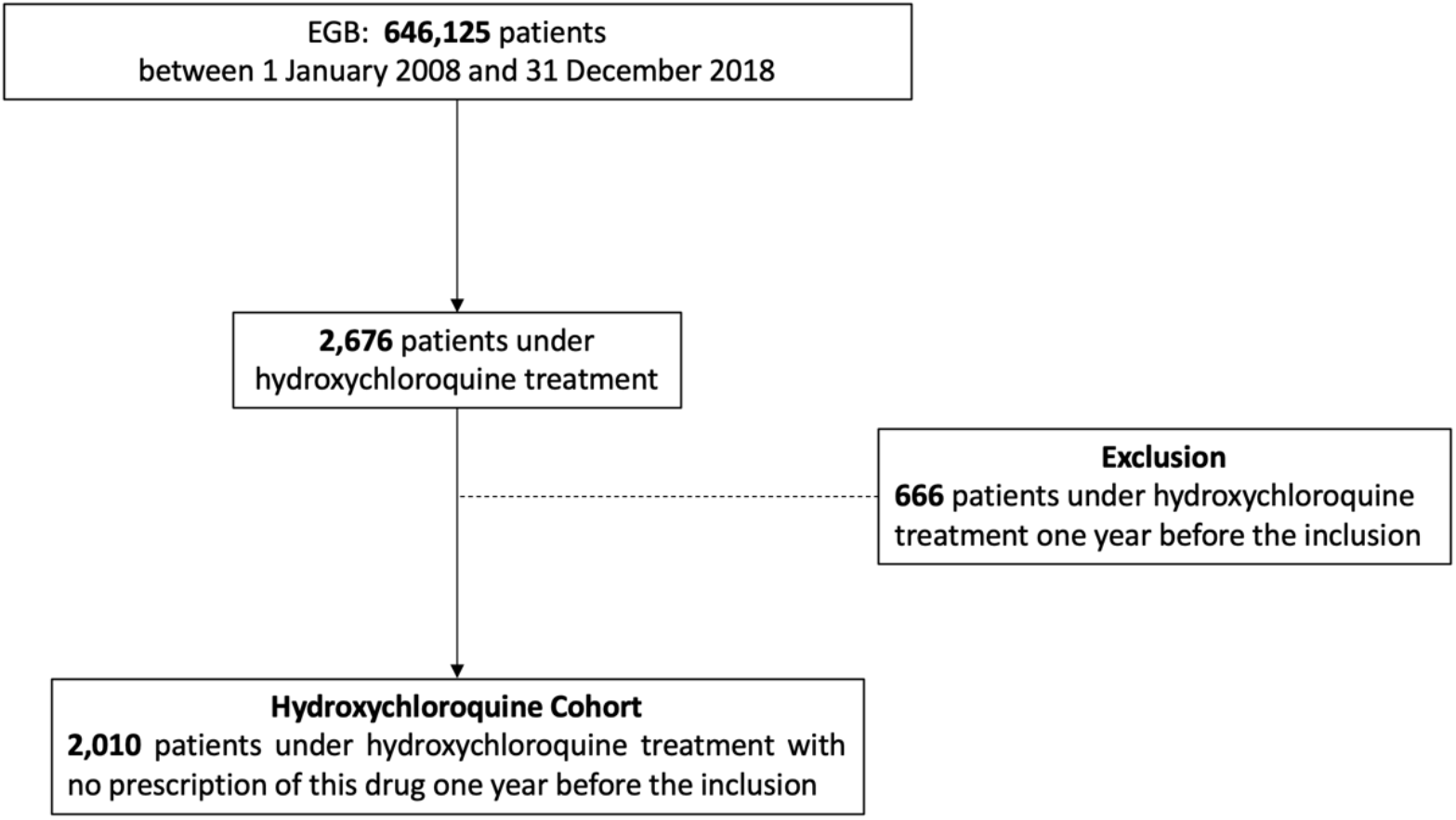
Population flowchart of hydroxychloroquine cohort.

All the parameters that were used in our study and that can be modified for other studies are presented in Table 1.

**Table 1:** Parameter set-up.

| Parameter | Set-up in our study | Explanation |
| --- | --- | --- |
| **Drug of interest** | Hydroxychloroquine | drug for which we want to know all co-prescriptions after exposure |
| **Study period** | 01/01/2008 to 12/31/2018 | study start and stop |
| **Time period** | month | time interval between start and stop |
| **Time window** | 24 months | potential exposure period before the event |
| **Covariate** | age, sex, and severity | covariate to fit the model |
| **Severity covariate** | value = 1 if the patient declared a chronic disease (lupus or rheumatoid polyarthritis) and/or received immunosuppressive treatment | immunosuppressive treatments are indicated for polyarthritis, which is an HCQ indication. We include other information available on disease severity for this study. |
| **Bootstraps** | 1,000 | repeat on sample of the WCE model application |

The EGB database contains only anonymized data and its access is legally authorized without having to receive authorization from the national data protection agency (CNIL). The study protocol was submitted to the appropriate INSERM and CNAMTS entities, as legally required. In addition, the study was approved by our Institutional Review Board (CER-APHP CENTRE, IRB n°20180603).

## RESULTS

The HCQ cohort contains 2,010 patients (n_women_ = 1,577, 78%), with 386 different ATC classes. The average age at the time of the first prescription of HCQ was 54.8 (sd = 16.2). Within the cohort, 1,045 (52%) patients had certification (received full reimbursement) for a chronic disease. Among them, 12% (n = 240) each had vasculitis, systemic lupus erythematosus, or systemic scleroderma, 11% (n = 214) rheumatoid arthritis, and 6% (n = 122) each malignant neoplasm or malignant disease of the lymphatic or hematopoietic tissue. The average duration of follow-up was > 10 years (Table 2). However, there was an association between age and HCQ exposure (P < 0.001 by linear regression analysis), as well as between sex and HCQ (P < 0.001 by the Wilcoxon test). There was also a strong association between HCQ exposure and severity (P < 0.001 by the Wilcoxon test).

**Table 2.** Population characteristics.

| Description | Hydroxychloroquine Cohort |
| --- | --- |
| **n (patients)** | 2,010 |
| number of women (%) | 1,577 (78%) |
| mean age (IQR) | 54.8 (23) |
| number of severe patients (%) | 701 (35%) |
| mean exposure, in months (sd) | 9.5 (16.1) |
| average follow-up, in years (sd) | 10.3 (1.3) |
| number of different ATC classes | 386 |
| **Number of patients with a chronic disease** | 1,045 (52%) |
| vasculitis, systemic lupus erythematosus, systemic scleroderma | 240 (12%) |
| rheumatoid arthritis | 214 (11%) |
| malignant neoplasm, malignant disease of the lymphatic or hematopoietic tissue | 122 (6%) |

The WCE-AI pipeline enabled the identification of nine ATC classes associated with the prescription of HCQ, of which five were borderline significant. The highest risk ratios were obtained for hydrocortisone (HR = 3.96 [1.66-7.55]), alendronic acid and cholecalciferol (HR = 3.24 [1.22-7.36]), 2.73 [1.03-6.13], valsartan (HR = 2.73 [1.03-6.13]), and chlormadinone (HR = 2.65 [1.16-4.76]) (Figure 5 & Appendix 1).

**Figure 5.**
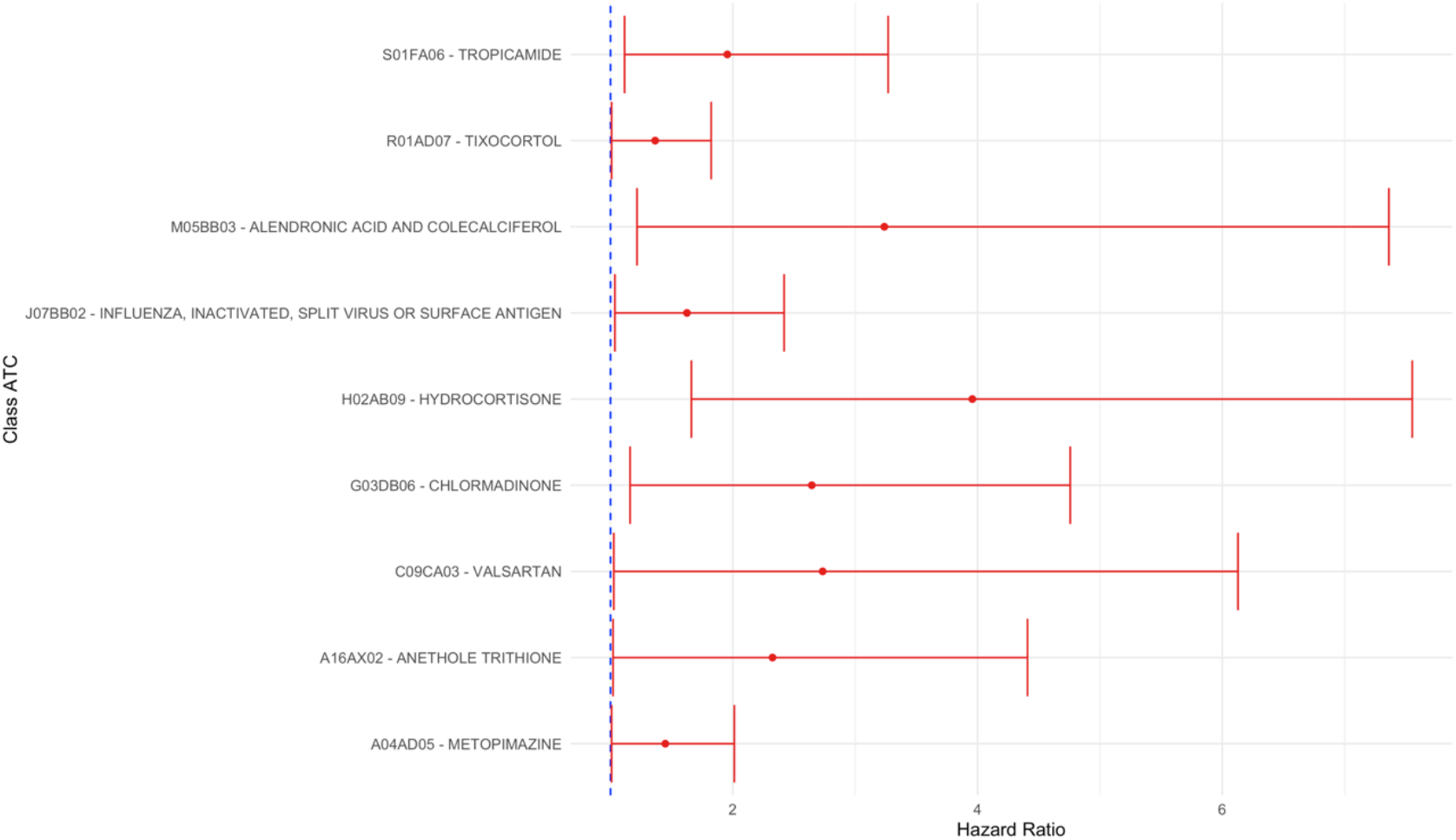
Forest plot of the effect of hydroxychloroquine on its co-prescription with the WCE-AI model.

The SCCO competitor method identified eight (81 before Bonferroni adjustment of p-values) ATC classes associated with HCQ prescription with control and risk periods of three months, 40 (162 before Bonferroni adjustment of p-values) for those of six months, and 67 (210 before Bonferroni adjustment of p-values) for those of nine months. The top 10 odds ratios per period are presented in Appendix 2.

In total, 0, 2, and 3 ATC classes were common between the WCE-AI and SCCO methods for the periods of 3, 6, and 9 months, respectively: anethole trithione and tropicamide (common to WCE-AI, SCCO 6 months and SCCO 9 months) and tixocortol (common to WCE-AI and SCCO 9 months).

## DISCUSSION

We report a new data-driven pipeline applied to HCQ on a cohort of 2,010 patients. This pipeline allowed us to identify nine ATC classes linked to HCQ, with the highest being alendronic acid and cholecalciferol, which are prescribed for osteoporosis.

Among the significant results we obtained, tropicamide, which is given in eye examinations for diagnostic purposes, is particularly relevant. Indeed, it is known that HCQ causes retinopathies and this ocular condition is monitored annually in people under such treatment (26). In addition, we identified a relevant signal for metopimazine, an antiemetic, with nausea being a frequently reported HCQ side-effect (27).

We found three drugs with a significant HR, as well as a significant severity coefficient: anethole trithione, a bile secretion-stimulating drug that restores salivation and relieves the discomfort of dry mouth in Sjögren’s syndrome, often associated with lupus (28); hydrocortisone, a glucocorticoid often used for withdrawal from more potent corticosteroids in the treatment of lupus or rheumatoid arthritis; and chlormadinone, a progestin macro-pill recommended as a contraceptive method for women with lupus (29,30). These three drugs, with a significant severity coefficient, are related to the management of the disease treated by HCQ and should not be considered as potential side effects.

Flu vaccine is also a co-prescription of HCQ. Indeed, influenza vaccination is a recommendation for patients with autoimmune disease who receive potentially immunosuppressive treatments (31).

Alendronic acid and cholecalciferol are also a co-prescription of HCQ for optimal management of patients with lupus or rheumatoid arthritis, mostly post-menopausal women, by internists or rheumatologists. In the literature, lupus and menopause are found to be two factors that favor osteoporosis (32,33). We believe that alendronic acid and cholecalciferol are not directly associated with HCQ but rather with the underlying disease.

Tixocortol is a glucocorticoid used as a nasal spray to treat allergic rhinitis. In the literature, there is a link between autoimmune diseases, such as lupus and allergic rhinitis. This link can be explained by the mechanism of these two diseases, which involves immune dysregulation and an increase in inflammatory mediators (34–38).

We did not find any clinically satisfactory explanation for the association with valsartan, an angiotensin II receptor blocker. It is know that HCQ can cause short-term conduction disorders (bundle or atrioventricular block) and, less frequently, long-term morphological abnormalities in heart tissue (myocardial hypertrophy) (39). Valsartan is given for high blood pressure and heart failure. It is impossible to know for which indication valsartan was prescribed, but because the signal is weak (borderline significant) and was not reproduced with the SCCO method, it may be a false positive signal due to multiple testing.

This pipeline did not detect any drugs that could reflect an adverse dermatological effect, even though it has been described in the literature (40).

The results of the SCCO method are difficult to interpret. This method is unable to separate ATC classes that are related to the course of the underlying disease (lupus or rheumatoid arthritis) from those that may represent a side effect.

### Study strengths and limitations

A major strength of this study was the use of a population-wide claims database based on the national public healthcare system, making the database population representative of the entire country population (41). The EGB is a particularly suitable basis for post-marketing authorization safety studies to assess the occurrence of rare adverse events (24,25,42–44).

Our data-driven pipeline is based on a self-controlled method, thus avoiding the usual biases of methods such as case-control (45) or propensity score (46) based approaches. In addition, we used exact temporal patterns to detect the broad spectrum of potential side effects of drugs and not only candidate side effects based on *a priori* hypotheses. Our method, with the exact temporal pattern of drug exposure, can identify short- and long-term side effects (16) and provides the possibility to adjust on covariates.

Our study also had several limitations. Claims databases are primarily used for healthcare reimbursement purposes and we assume that the reimbursed drugs are consumed by the patient. These databases are extensive and can pose problems in terms of the statistical significance of the results and their interpretation. We only used drugs reimbursed outside the hospital. It may be informative to supplement this data source with hospital prescriptions. Another limitation is that our method only considers the co-prescription of drugs as the source of adverse drug reaction signals. Indeed, certain adverse drug effects do not require the prescription of another drug. For example, prolongation of the QT interval after exposure to HCQ cannot be detected by our method because the treatment for prolongation of the QT interval is the discontinuation of HCQ. Similarly, our method will not be able to detect fatal side effects or ototoxicity of HCQ (47).

In conclusion, we present a data-driven pipeline that provides a broad picture of the side effects of a given drug. Applied to HCQ, our pipeline allowed us to show that the adverse effects of HCQ are essentially known and controlled by health professionals.

This pipeline could not highlight all the known side effects of HCQ, such as cardiac toxicity, because it relies on a single source of data, drug prescriptions. It would be useful to pursue the development of this pipeline by bringing in other sources of data.

## Data Availability

Data not available

## ACKNOWLEDGEMENTS

The authors gratefully acknowledge the contribution of Marie Jamin and Brigitte Sabatier for their valued comments on this manuscript.

## AUTHOR CONTRIBUTIONS

P.S., M.W., J.P., N.D., and AS.J. wrote the manuscript. P.S., N.D., and AS.J. designed the research. P.S. performed the research. P.S., M.W., J.P., N.D., and AS.J. analyzed the data.

---

**Appendix 1.** ATC Class associated with hydroxychloroquine prescription in the WCE-AI model.

| Class ATC | n | HR [CI<sub>5%</sub>] | Age coefficient [CI<sub>5%</sub>] | Sex coefficient [CI<sub>5%</sub>] | Severity coefficient [CI<sub>5%</sub>] |
| --- | --- | --- | --- | --- | --- |
| **A04AD05 - METOPIMAZINE** | 747 | 1.45 [1.01-2.01] | 0.99 [0.99-0.99] | 1.42 [1.19-1.72] | 0.92 [0.79-1.05] |
| **A16AX02 - ANETHOLE TRITHIONE** | 138 | 2.32 [1.02-4.41] | 1.02 [1.01-1.04] | 1.78[1.12-3.76] | 1.58 [1.08-2.33] |
| **S01FA06 - TROPICAMIDE** | 254 | 1.96 [1.12-3.27] | 1.05 [1.03-1.06] | 0.48 [0.29-0.81] | 1.47 [0.90-2.46] |
| **M05BB03 - ALENDRONIC ACID AND COLECALCIFEROL** | 71 | 3.24 [1.22-7.36] | 0.97 [0.96-0.97] | 27.66 [8.24-95630273.85] | 1.43 [0.95-2.04] |
| **H02AB09 - HYDROCORTISONE** | 69 | 3.96 [1.66-7.55] | 1.01 [1.0-1.03] | 0.90 [0.52-1.94] | 3.07 [1.86-5.25] |
| **G03DB06 - CHLORMADINONE** | 142 | 2.65 [1.16-4.76] | 1.03 [1.03-1.04] | 0.84 [0.69-1.05] | 1.42 [1.16-1.72] |
| **C09CA03 - VALSARTAN** | 79 | 2.73 [1.03-6.13] | 1.04 [1.03-1.06] | 1.61 [0.80-4.54] | 1.28 [0.77-2.11] |
| **J07BB02 - INFLUENZA, INACTIVATED, SPLIT VIRUS OR SURFACE ANTIGEN** | 545 | 1.63 [1.04-2.42] | 0.99 [0.99-1.0] | 1.20 [1.05-1.40] | 1.01 [0.89-1.13] |
| **R01AD07 - TIXOCORTOL** | 927 | 1.37 [1.01-1.82] | 1.05 [1.04-1.05] | 0.90 [0.68-1.25] | 1.09 [0.83-1.40] |

**Appendix 2.** Top 10 ATC classes associated with hydroxychloroquine prescription in the SCCO model after Bonferroni adjustment of p values

| Top 10 class ATC, SCCO model with periods (risk and control) of 3 months | OR [CI<sub>5%</sub>] |
| --- | --- |
| **M01AH01 - CELECOXIB** | 3.88 [2.04-7.36] |
| **L04AX03 - METHOTREXATE** | 2.93 [1.83-4.7] |
| **S01XA20 - ARTIFICIAL TEARS AND OTHER INDIFFERENT PREPARATIONS** | 2.6 [1.77-3.83] |
| **N05BA12 - ALPRAZOLAM** | 2.51 [1.65-3.81] |
| **H02AB07 - PREDNISONE** | 2.27 [1.57-3.29] |
| **A07XA04 - RACECADOTRIL** | 2.24 [1.51-3.32] |
| **H02AB06 - PREDNISOLONE** | 1.97 [1.41-2.76] |
| **A11CC05 - COLECALCIFEROL** | 1.81 [1.36-2.4] |

| Top 10 class ATC, SCCO model with periods (risk and control) of 6 months | OR [CI<sub>5%</sub>] |
| --- | --- |
| **M01AH01 - CELECOXIB** | 6.46 [2.7-15.46] |
| **A12AA04 - CALCIUM CARBONATE** | 5.04 [2.52-10.09] |
| **L04AX03 - METHOTREXATE** | 4.97 [3.16-7.83] |
| **A16AX02 - ANETHOLE TRITHIONE** | 4.04 [2.02-8.07] |
| **H02AB07 - PREDNISONE** | 3.8 [2.73-5.29] |
| **M01AH01 - CELECOXIB** | 3.34 [1.92-5.8] |
| **R06AX13 - LORATADINE** | 3.32 [1.85-5.96] |
| **J02AA01 - AMPHOTERICINE B** | 3.26 [1.88-5.64] |
| **A02BC04 - RABEPRazole** | 3.25 [1.86-5.7] |
| **J01AA02 - DOXYCYCLINE** | 3.23 [2.04-5.13] |

| Top 10 class ATC, SCCO model with periods (risk and control) of 9 months | OR [CI<sub>5%</sub>] |
| --- | --- |
| **M01AH01 - CELECOXIB** | 6.74 [4.36-10.43] |
| **P01AB02 - TINIDAZOLE** | 6.4 [2.48-16.54] |
| **A16AX02 - ANETHOLE TRITHIONE** | 5.48 [2.71-11.08] |
| **L01BA01 - METHOTREXATE** | 4.65 [2.74-7.89] |
| **H02AB07 - PREDNISONE** | 4.4 [3.21-6.03] |
| **A12AA04 - CALCIUM CARBONATE** | 4.14 [2.22-7.72] |
| **B03BB01 - FOLIC ACID** | 3.74 [2.72-5.14] |
| **A01AD11 - VARIOUS** | 3.63 [2.12-6.2] |
| **N06AB05 - PAROXETINE** | 3.6 [2.05-6.31] |
| **N02AA01 - MORPHINE** | 3.54 [1.99-6.3] |

